# Methotrexate and Prednisolone compared to placebo and prednisolone in the treatment of Erythema Nodosum Leprosum - an international multicentre, double-blind randomised controlled clinical trial - MaPs in ENL

**DOI:** 10.64898/2026.05.19.26353561

**Authors:** Barbara de Barros, Farha Sultana, Neeta Maximus, Vivek V. Pai, Anju Wakade, Bhagyashree Bhame, Bishwanath Acharya, Abdulnaser Hamza, Alemtsehay Getachew, Medhi Denisa Alinda, M. Yulianto Listiawan, Shimelis N Doni, Deanna A. Hagge, Indra Napit, Mahesh Shah, Joydeepa Darlong, Peter Nicholls, Bernd Genser, Saba M. Lambert, Diana N. J. Lockwood, Stephen L. Walker, the Erythema Nodosum Leprosum International Study Group

## Abstract

**Background:** Erythema nodosum leprosum (ENL) is a severe inflammatory complication of leprosy that often requires prolonged corticosteroid therapy which is associated with adverse effects. Methotrexate is an affordable immunomodulatory agent with limited evidence for its use in ENL treatment. We evaluated whether weekly oral methotrexate in additional to prednisolone reduces the need for additional prednisolone in adults with severe ENL.

**Methods and Findings:** We performed an international, multicentre, double-blind, randomised, placebo-controlled trial conducted at five leprosy referral centres in Ethiopia, India, Indonesia, and Nepal. Adults aged 18–60 years with severe ENL were randomised to receive oral methotrexate and prednisolone, or matching placebo and prednisolone. All participants received an identical prednisolone regime over 20 weeks and were followed for 60 weeks. The primary outcome was time to first ENL flare requiring additional prednisolone, assessed over 24 and 48 weeks. Between January 2023 and June 2024, 231 individuals were screened and 137 were randomised (68 methotrexate and prednisolone; 69 placebo and prednisolone). By 24 weeks, 85/137 (62.0%) participants experienced an ENL flare requiring additional prednisolone; the adjusted hazard ratio (HR) for methotrexate versus placebo was 0.98 (95% CI 0.62–1.54). By 48 weeks, 102/137 (74.5%) experienced an ENL flare; adjusted HR 0.95 (95% CI 0.62–1.43). Secondary outcomes were similar: methotrexate did not reduce ENL severity at first flare, flare frequency, or severity of subsequent flares. Health-related quality of life improved substantially in both groups with no evidence of a differential treatment effect. Methotrexate was generally well tolerated. The trial was registered at ClinicalTrials.gov (NCT03775460).

**Conclusions:** Oral methotrexate added to prednisolone did not reduce the requirement for additional prednisolone or delay ENL flares compared to placebo and prednisolone, and our study does not support the use of methotrexate for severe ENL.

**Author’s Summary:** *Why was this study done?:*

- Erythema nodosum leprosum (ENL) is a painful, relapsing inflammatory complication of leprosy that often requires prolonged courses of oral corticosteroids.
- Long-term corticosteroids can cause serious adverse effects including mortality in some settings
- Thalidomide is effective but is not accessible in many endemic countries.
- Methotrexate treatment is inexpensive and widely available, but high-quality evidence for its use in ENL is lacking.

*What did the researchers do and find?:*

- We conducted an international, multicentre, double-blind randomised trial in four countries, enrolling 137 adults with severe ENL.
- Participants received either weekly oral methotrexate or identical placebo tablets, and 20-week course of oral prednisolone
- Methotrexate did not reduce the likelihood of ENL flares needing additional prednisolone at 24 or 48 weeks, nor did it reduce flare severity or overall flare burden.
- Quality of life improved markedly but with no clear difference between treatment groups.
- Methotrexate was generally well tolerated

*What do these findings mean?:*

- Adding methotrexate to prednisolone does not provide clinically meaningful corticosteroid-sparing benefit for severe ENL.
- New therapies for severe ENL are required because even with high quality care corticosteroids are required at high doses for prolonged periods in most people

## INTRODUCTION

Leprosy is a chronic granulomatous disease caused by *Mycobacterium leprae* and *Mycobacterium lepromatosis* [1]. The World Health Organization (WHO) reported 172,717 new cases in 2024, with nearly 80% diagnosed in India, Brazil and Indonesia [2].

Leprosy is complicated by immune-mediated inflammatory episodes known as leprosy reactions [1] which may occur before, during or after successful completion of anti-microbial WHO recommended multi-drug therapy (MDT). Leprosy reactions affect more than 60% of Individuals with multibacillary leprosy [1,3,4]. Erythema nodosum leprosum (ENL)or leprosy Type 2 reaction, is a severe reaction which affects up to 50% of individuals with lepromatous leprosy (LL) and up to 10% of those with borderline lepromatous (BL) leprosy. A bacterial index of four or more is a risk factor for developing ENL [3,5,6]. ENL is characterised by episodes of painful erythematous cutaneous nodules accompanied by systemic symptoms, including fever, arthritis, neuritis and orchitis and typically affects economically active adults in their 20s and 30s [7]. Individuals with ENL experience disability, reduced health-related quality of life (HRQoL) and catastrophic household costs [8–11].

ENL involves complement activation and neutrophil infiltration of tissues, with elevated plasma levels of pro-inflammatory cytokines including tumour necrosis factor alfa (TNF-α), interleukins (IL-2, IL-6, IL-10, IL-8, IL-12) and interferon gamma (INF-γ) [12–15].

ENL is often chronic requiring prolonged courses of high dose oral corticosteroid therapy which WHO the recommend as first-line treatment [16]. Corticosteroid therapy in patients with ENL is associated with adverse effects and significant mortality in some settings [17–19]. Thalidomide is highly effective for ENL however, its use is restricted in many endemic countries due to concerns regarding teratogenicity [18,20–22]. Consequently, there is a critical unmet need for safe, affordable and effective corticosteroids-sparing therapies for ENL.

Methotrexate inhibits the enzyme dihydrofolate reductase, essential for the synthesis of purines and pyrimidines, and is effective in a range of chronic inflammatory diseases, including rheumatoid arthritis and psoriasis [23–25]. Methotrexate modulates cytokine production including TNF-α, IL-6 and INF-γ and exerts anti-inflammatory effects at low doses and is not associated with the broad cytotoxic immunosuppression seen with many other immunosuppressive agents [24]. Oral or subcutaneous methotrexate at doses usually between 7.5mg to 25mg weekly is widely used in both adult and paediatric dermatology practice and so is familiar to dermatologists and leprologists. Methotrexate is affordable and widely available in low-and middle-income settings [26,27]. Methotrexate has been used for ENL in patients with chronic or refractory disease in small observational studies and case series. The only previous trial evaluating methotrexate in ENL was an open label, randomised trial which recruited 19 participants in Bangladesh [28], 10 participants received 7.5 mg of methotrexate weekly for 24 weeks and 12 weeks of prednisolone and the other nine received prednisolone for 24 weeks. There were no serious adverse effects.

Leprosy reactions, in particular ENL, are recognised as priority conditions in leprosy research and control strategies due to their frequency, severity and disproportionate contribution to morbidity, mortality, disability and health-related quality of life impact [16].

We wished to assess the efficacy and safety of weekly oral methotrexate treatment in adults with severe ENL and conducted a double-blind, randomised, placebo-controlled multi-centre trial-to determine whether methotrexate and prednisolone reduced the need for additional prednisolone compared with prednisolone alone.

## METHODS

### Trial design and conduct

The study methods, conduct and analysis were described previously in the published protocol and statistical analysis plan, are available in the Supplementary File 1. The trial was designed and conducted by the ENLIST Group [29]. The London School of Hygiene & Tropical Medicine (LSHTM) was the sponsor. The funders and donors had no role in the design or conduct of the trial. All authors vouch for the completeness and accuracy of the data and for the fidelity of the trial to the protocol.

### Trial sites and participants

Individuals diagnosed with severe ENL, ENLIST ENL Severity Scale (EESS) score of nine or more [30], between 18 and 60 years of age who gave informed consent were recruited from five leprosy referral facilities in four countries: the ALERT Hospital, Addis Ababa, Ethiopia; The Leprosy Mission Trust India Barabanki Hospital, Barabanki and Bombay Leprosy Project, Mumbai, India; Dr. Soetomo Hospital, Surabaya, Indonesia and The Leprosy Mission Nepal Anandaban Hospital, Kathmandu, Nepal. Exclusion criteria were: ENL present for more than 4 years, pregnancy, breastfeeding, hepatitis B virus or C virus or HIV infection, tuberculosis, pulmonary fibrosis or any contra-indication to methotrexate [31].

### Definitions of ENL

ENL was defined as occurring when an individual with BL leprosy or LL developed 10 or more tender papular and/or nodular skin. Participants were classified as having one of three clinical patterns of ENL acute or recurrent or chronic [31].

### Trial oversight

Written informed consent to participate in the trial was obtained from all participants. The study was approved by local and national ethics and regulatory authorities in all sites and from LSHTM in the United Kingdom. An independent data and safety monitoring board (DSMB) reviewed data every six months during the trial.

### Randomisation and allocation concealment

Participants were randomly assigned in a 1:1 ratio to receive either methotrexate or placebo. The randomisation list was computer-generated in blocks of six. Methotrexate 2.5 mg tablets or identical placebo tablets were prepared in individualised identical packets by a specialist company. Intervention packets for each participant were packed in a box identified by a serial number determined by the randomisation list. Each box contained 16 packets with either methotrexate or placebo tablets and boxes were distributed to centres maintaining the serial numbering and allocated to participants in order by serial number. Prednisolone tablets were procured by each centre from their usual suppliers. Each centre had a pharmacist responsible for the study medication, storage and distribution.

To verify the integrity of the investigational products and the effectiveness of allocation concealment, independent drug quality testing was undertaken during the trial. Methotrexate tablets and placebo tablets samples drawn from unused participant boxes were analysed using a validated high-performance liquid chromatography method. Methotrexate was detected exclusively in samples allocated to the active treatment arm, while placebo samples contained no active pharmaceutical ingredients. Prednisolone tablets from each participating centre were tested. These analyses confirmed the chemical identity and consistency of the study medications and provided independent validation of the randomisation and allocation concealment procedures implemented in the trial.

### Trial interventions

Participants in the intervention arm received an initial dose of oral methotrexate of 10mg, then 15mg seven days later, participants weighing less than 60kg continued to receive 15mg of methotrexate weekly for the duration of the intervention. Individuals weighing 60kg or more had a weekly dose of methotrexate 20mg from dose 9 onwards. At week 48, the methotrexate was reduced to 10mg weekly for all participants for 2 weeks followed by 5mg weekly for 2 weeks and then stopped. In the control arm, participants received an identical number of placebo tablets depending on their weight. The dose of methotrexate was chosen in accordance with safe and effective prescribing of methotrexate for skin disease [27].

Trial participants in both arms received an identical regime of oral prednisolone for 20 weeks. The initial dose was 40mg daily, reducing to zero over 20 weeks (Table 1).

**Table 1:** Intervention by allocation group. Participants weighing less than 60kg continued to receive 15mg of methotrexate weekly and individuals weighing 60kg or more received 20mg from week 8.

| Time from enrolment | Methotrexate and Prednisolone group |  | Placebo and Prednisolone group |  |
| --- | --- | --- | --- | --- |
| Week | Methotrexate (mg/WEEK)<br>(Methotrexate tablets 2.5 mg) | Prednisolone (mg/day) | Prednisolone (mg/day) | Placebo<br>(Number of tablets/WEEK) |
| 0 | 10 | 40 | 40 | 4 |
| 1 | 15 | 40 | 40 | 6 |
| 2 | 15 | 35 | 35 | 6 |
| 3 | 15 | 35 | 35 | 6 |
| 4 | 15 | 30 | 30 | 6 |
| 5 | 15 | 30 | 30 | 6 |
| 6 | 15 | 25 | 25 | 6 |
| 7 | 15 | 25 | 25 | 6 |
| 8 | 15/20 | 20 | 20 | 6/8 |
| 9 | 15/20 | 20 | 20 | 6/8 |
| 10 | 15/20 | 20 | 20 | 6/8 |
| 11 | 15/20 | 20 | 20 | 6/8 |
| 12 | 15/20 | 15 | 15 | 6/8 |
| 13 | 15/20 | 15 | 15 | 6/8 |
| 14 | 15/20 | 15 | 15 | 6/8 |
| 15 | 15/20 | 15 | 15 | 6/8 |
| 16 | 15/20 | 10 | 10 | 6/8 |
| 17 | 15/20 | 10 | 10 | 6/8 |
| 18 | 15/20 | 5 | 5 | 6/8 |
| 19 | 15/20 | 5 | 5 | 6/8 |
| 20-47 | 15/20 | 0 | 0 | 6/8 |
| 48 | 10 | 0 | 0 | 4 |
| 49 | 10 | 0 | 0 | 4 |
| 50 | 5 | 0 | 0 | 2 |
| 51 | 5 | 0 | 0 | 2 |
| 52-59 | 0 | 0 | 0 | 0 |

### Medication

All participants received folic acid 5 mg daily to reduce the risk of methotrexate-associated toxicity [27]. In addition, gastric and bone protection were prescribed concomitantly with prednisolone, in accordance with standard practice at each study site. Ivermectin 200 mg/kg body weight was administered at enrolment and repeated every six months during follow-up [31].

### Outcomes

#### Primary outcomes

The primary outcome was the requirement of additional prednisolone by participants. These measurements were chosen because the requirement for additional prednisolone is a proxy marker of ENL deterioration and treatment failure. We chose two different evaluation periods: i) only counting events during the first 24 weeks and ii) all events during the first 48 weeks.

#### Secondary outcomes

The secondary outcomes to be assessed by comparisons between treatment arms were: (1) change in EESS score between enrolment and first flare of ENL, (2) the proportion of individuals who did not require prednisolone at 60 weeks, (3) the number of flares of ENL per individual requiring additional prednisolone up to 60 weeks, (4) severity of flares of ENL, (5) time to first flare after enrolment, (6) changes in participant-reported HRQoL at 48 and 60 weeks, and (7) the proportion of individuals with treatment related adverse events (AEs).

### Assessment and investigations

Participants were followed for 60 weeks and assessed at 20 scheduled visits, including at enrolment; weekly visits during the first 2 weeks; fortnightly visits until week 12; and four-weekly visits from week 16 to week 60. Additional unscheduled visits were permitted as clinically indicated for the assessment and management of ENL flares or other intercurrent events. At each study visit, a structured clinical history was obtained, and a comprehensive clinical examination was performed, including nerve function assessment and evaluation of EESS score.

The EESS is a 10-item validated severity scale. Scores can range from 0 to 30, higher scores indicate more severe ENL. A score of 9 or more is classified as severe ENL and a 5-point difference is clinically meaningful [30].

Laboratory assessments included full blood count, renal and liver function tests, and glucose measurement, with urinary pregnancy testing performed for women of reproductive age. At screening, all participants had HIV, hepatitis B virus (surface antigen and core antibody), and hepatitis C virus serologies performed. A chest radiograph was obtained prior to enrolment.

### Outcome measures

The primary outcome event was ENL flare requiring additional prednisolone, which was defined as the presence of ENL-related symptoms and/or signs meeting at least one of the following criteria: an increase in the EESS score to nine or more; an increase in EESS score of five or more from the previous assessment; orchitis not responding to conservative management; or iritis not responding to topical corticosteroids and/or mydriatic therapy.

Secondary outcomes included the severity of ENL at recurrence, the frequency of treatment-related adverse events, and changes in HRQoL over the follow-up period. HRQoL was assessed using the appropriate validated language versions of the Dermatology Life Quality Index (DLQI)[32] and the 36-Item Short Form Health Survey (SF-36)[33] administered at enrolment and weeks 24, 48, and 60. The DLQI evaluates the impact of skin disease on quality of life using a total score ranging from 0 to 30, with higher scores indicating greater impairment; scores of more than 10 represent a very large or extremely large effect on HRQoL [32]. The SF-36 is a generic measure of HRQoL assessing eight domains of physical and mental health, with domain scores ranging from 0 to 100, where higher scores indicate better perceived health status [33].

Adverse events were assessed at each scheduled study visit using standardised case report forms. Anticipated adverse events related to methotrexate and prednisolone were actively monitored, supported by routine laboratory investigations performed according to the study protocol.

### Sample size calculation

The sample size was calculated prior to trial initiation. To detect a clinically meaningful difference with 90% power at a two-sided α of 0.05, 416 participants were required, including 20% loss to follow-up.

### Data management

Each participant was assigned a unique study identification number, and all data were recorded and analysed with this unique identification number. Data were entered into a Good Clinical Practice compliant, trial-specific database [Research Electronic Data Capture (RedCap), Nashville, TN, USA].

### Statistical analysis

Owing to delays related to the COVID-19 pandemic, withdrawal of participating sites and periods of civil unrest the recruitment target was not achieved so the statistical analysis plan was revised prior to unblinding. The revised analyses emphasised effect sizes with 95% confidence intervals rather than formal hypothesis testing. Acute and recurrent or chronic ENL were analysed within a single combined analysis population to maximise statistical power while retaining adjustment for ENL phenotype in multivariable models. This approach was prespecified in the revised analysis plan and reflects the shared clinical objective of reducing corticosteroid-requiring ENL flares across ENL types (acute and recurrent or chronic).

The primary objective of the statistical analyses was to evaluate the efficacy and safety of methotrexate in the treatment of adult participants with acute and recurrent or chronic ENL. The primary efficacy analyses were conducted in the intention-to-treat population. Safety analyses were conducted in the safety population defined as all participants who received at least one dose of study medication. All statistical analyses were conducted with treatment allocation concealed.

The primary outcome, ENL flare requiring prednisolone, was analysed by comparing the proportion of participants who required additional prednisolone and a time-to-event analysis considering the event (first use of prednisolone for two different follow-up periods (24 and 48 weeks). Univariate (Kaplan-Meier curves) and multivariate time-to-event methods (Cox proportional hazards regression) were used to analyse time to first additional prednisolone. Adjusting variables for the Cox models were *a priori* specified including all available prognostic factors, including ENL type (acute vs recurrent or chronic), sex, age category, weight, EESS score at enrolment and WHO-MDT status. Proportional hazards assumption was assessed by diagnostic plots (log-log plot) and proportional hazards test.

Individuals that were lost to follow up were censored at last study visit. A sensitivity analysis was performed to ensure that all randomised participants contributed to the primary analyses. In addition, a per-protocol analysis was performed as a robustness check, restricted to participants without major protocol deviations, to assess the consistency of findings with the ITT analyses. Statistical analysis was conducted using STATA (StataCorp. *Stata Statistical Software: Release 18*. College Station, TX: StataCorp LLC, 2023).

### Ethics statement

The trial was conducted in accordance with the Helsinki Declaration as revised in 2024. Ethical approval was obtained from the LSHTM Research Ethics Committee (15762). Approval was obtained from Dr. Soetomo Hospital ethics committee, Indonesian Food and Drug Authority, Ethiopian Ministry of Education ethics committee, Ethiopian Food and Drug Authority,

Bombay Leprosy Project committee, the Leprosy Mission Trust India Ethics Committee, Nepal Health Research Council and Department of Drug Administration, Nepal Government.

This study was registered at www.clinicaltrials.gov (NCT03775460).

## RESULTS

### Trial population

Between 2^nd^ January 2023 and 29^th^ June 2024, 231 individuals with ENL were screened and 137 participants were enrolled and randomised across five sites in four countries. One hundred and sixteen participants were recruited in India, eleven in Nepal, five in Indonesia and five in Ethiopia. All randomised participants were included in the intention-to-treat population. One participant was excluded from adjusted regression analyses due to missing baseline covariate data, leaving 136 participants in adjusted Cox models.

**Figure 1:**
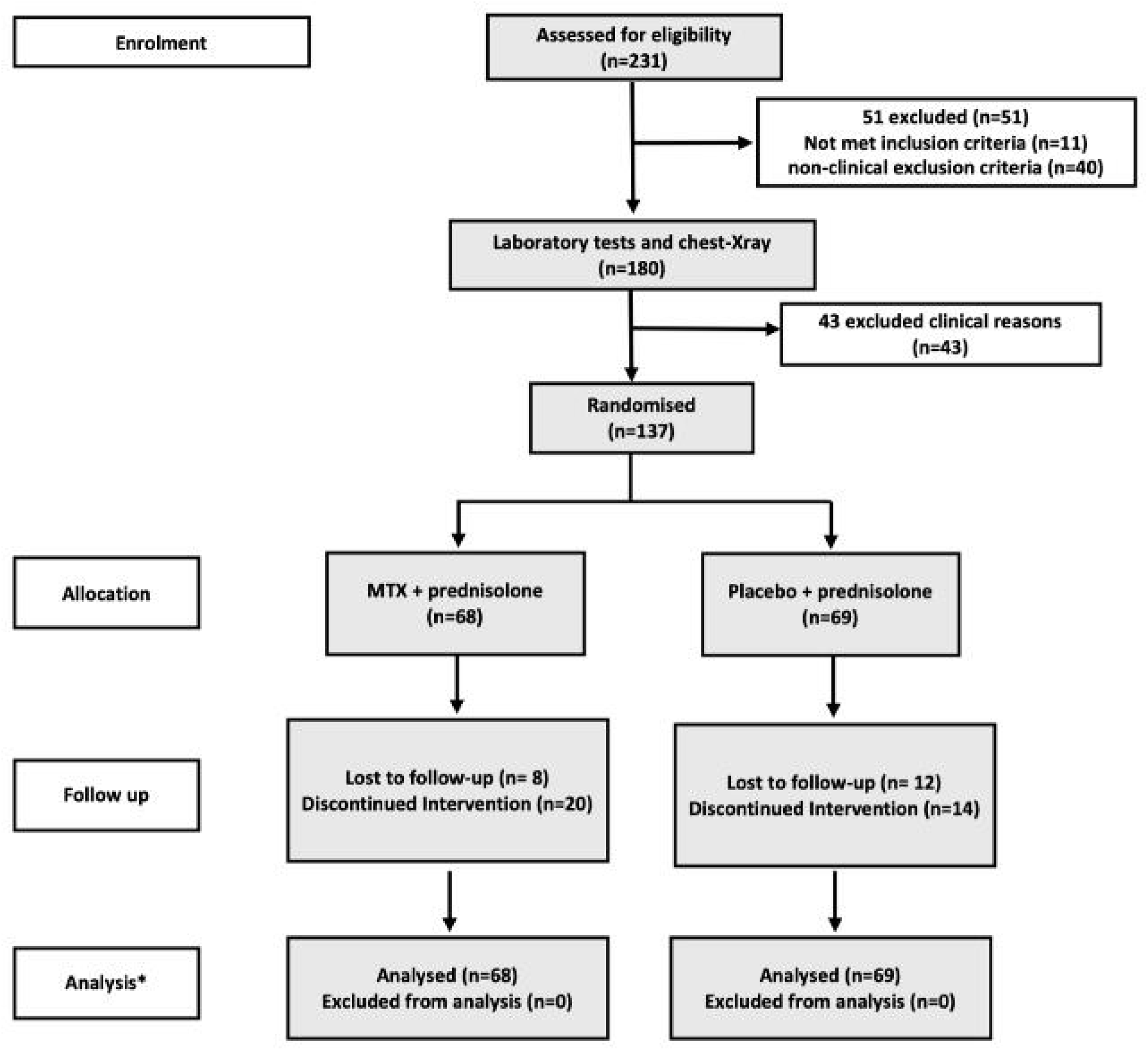
CONSORT flow diagram for MaPs in ENL with individuals randomised to either methotrexate (MTX) and prednisolone or placebo and prednisolone. * All participants randomised were included in the intention-to-treat population.

### Enrolment characteristics

Demographic and clinical characteristics at enrolment were well balanced between the two treatment arms (Table 2). Overall, 102/137 (74.5%) participants were male, with a median age of 33 years (IQR 16). Most participants had recurrent or chronic ENL (97/137; 70.8%), and the majority had LL (87.1%).

**Table 2:**
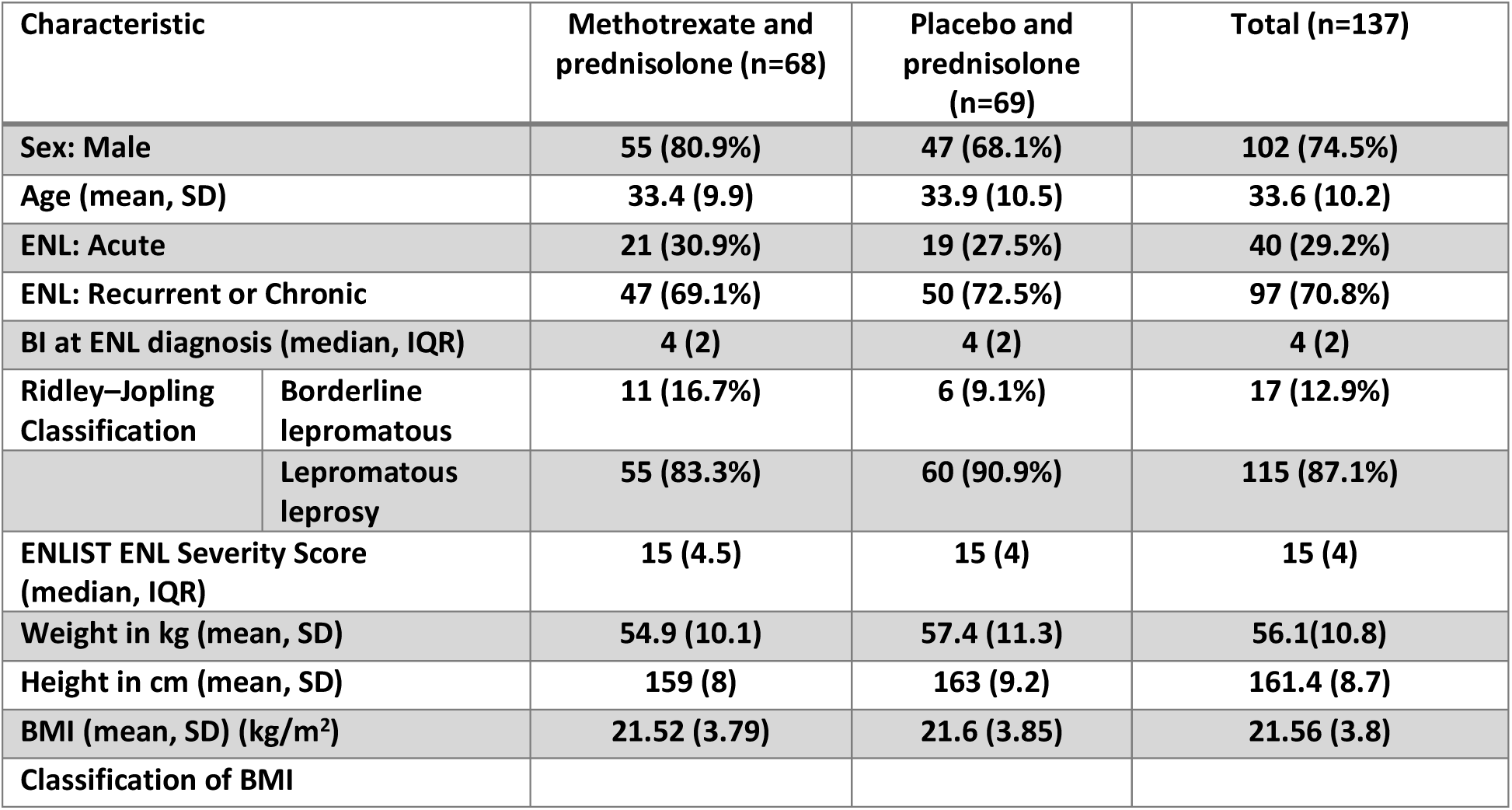
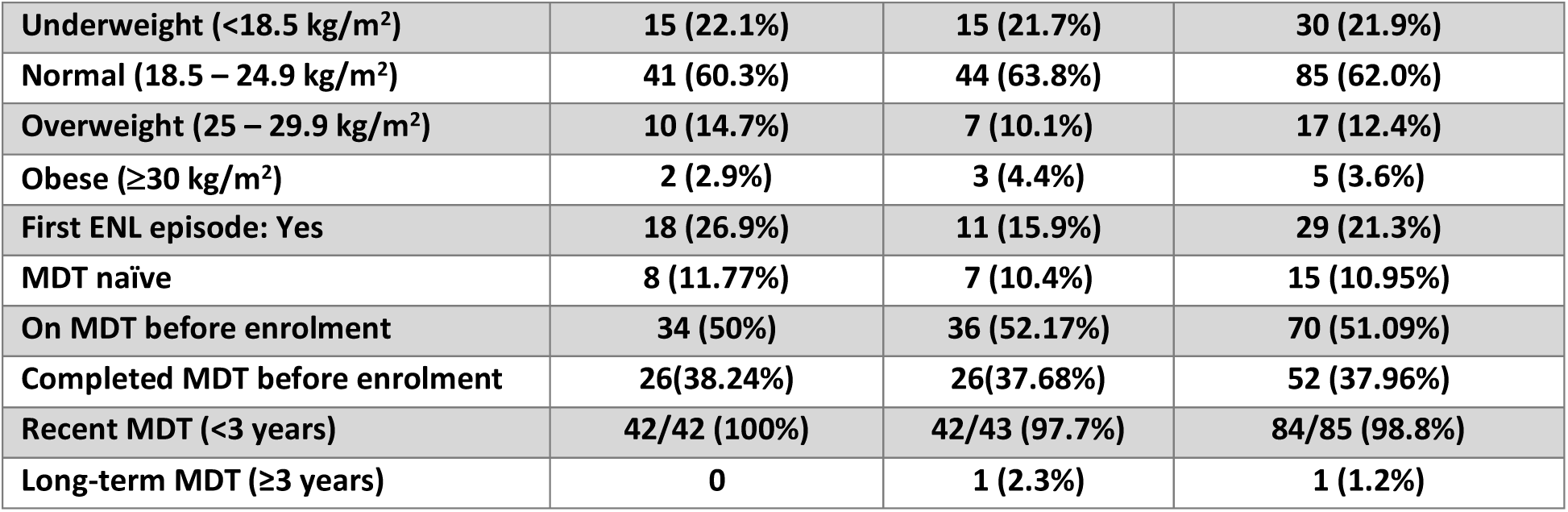
Characteristics of the intention-to-treat population at enrolment.

| Characteristic |  | Methotrexate and prednisolone (n=68) | Placebo and prednisolone (n=69) | Total (n=137) |
| --- | --- | --- | --- | --- |
| Sex: Male |  | 55 (80.9%) | 47 (68.1%) | 102 (74.5%) |
| Age (mean, SD) |  | 33.4 (9.9) | 33.9 (10.5) | 33.6 (10.2) |
| ENL: Acute |  | 21 (30.9%) | 19 (27.5%) | 40 (29.2%) |
| ENL: Recurrent or Chronic |  | 47 (69.1%) | 50 (72.5%) | 97 (70.8%) |
| BI at ENL diagnosis (median, IQR) |  | 4 (2) | 4 (2) | 4 (2) |
| Ridley–Jopling Classification | Borderline lepromatous | 11 (16.7%) | 6 (9.1%) | 17 (12.9%) |
|  | Lepromatous leprosy | 55 (83.3%) | 60 (90.9%) | 115 (87.1%) |
| ENLIST ENL Severity Score (median, IQR) |  | 15 (4.5) | 15 (4) | 15 (4) |
| Weight in kg (mean, SD) |  | 54.9 (10.1) | 57.4 (11.3) | 56.1(10.8) |
| Height in cm (mean, SD) |  | 159 (8) | 163 (9.2) | 161.4 (8.7) |
| BMI (mean, SD) (kg/m<sup>2</sup>) |  | 21.52 (3.79) | 21.6 (3.85) | 21.56 (3.8) |
| Classification of BMI |  |  |  |  |

|  |  |  |  |
| --- | --- | --- | --- |
| Underweight (<18.5 kg/m<sup>2</sup>) | 15 (22.1%) | 15 (21.7%) | 30 (21.9%) |
| Normal (18.5 – 24.9 kg/m<sup>2</sup>) | 41 (60.3%) | 44 (63.8%) | 85 (62.0%) |
| Overweight (25 – 29.9 kg/m<sup>2</sup>) | 10 (14.7%) | 7 (10.1%) | 17 (12.4%) |
| Obese (≥30 kg/m<sup>2</sup>) | 2 (2.9%) | 3 (4.4%) | 5 (3.6%) |
| First ENL episode: Yes | 18 (26.9%) | 11 (15.9%) | 29 (21.3%) |
| MDT naïve | 8 (11.77%) | 7 (10.4%) | 15 (10.95%) |
| On MDT before enrolment | 34 (50%) | 36 (52.17%) | 70 (51.09%) |
| Completed MDT before enrolment | 26(38.24%) | 26(37.68%) | 52 (37.96%) |
| Recent MDT (<3 years) | 42/42 (100%) | 42/43 (97.7%) | 84/85 (98.8%) |
| Long-term MDT (≥3 years) | 0 | 1 (2.3%) | 1 (1.2%) |

The median EESS score at enrolment was 15 (IQR 4) in both arms. Almost two-thirds of participants were receiving MDT at enrolment (62.5%), with nearly all having commenced MDT within the preceding three years. Nutritional status was comparable between arms, with a mean body mass index (BMI) of 21.56 kg/m² (SD 3.8).

### Primary outcome

#### Requirement for additional prednisolone at 24 weeks and 48 weeks

By 24 weeks, 85 of 137 participants (62%) had experienced at least one ENL flare requiring additional prednisolone. This occurred in 41 of 68 (60.3%) participants in the methotrexate and prednisolone group and in 44 of 69 (63.8%) participants in the placebo and prednisolone group. The Kaplan-Meier estimate of remaining free from ENL-related additional prednisolone at 24 weeks was 39.7% (95%CI 28.1-51) in the methotrexate and prednisolone group and 36.2% (95%CI 25.1-47.4) in the placebo and prednisolone group (Figure 2). In the adjusted Cox proportional hazard model, there was no evidence of a difference in the hazard of experiencing a first ENL flare requiring additional prednisolone between participants receiving methotrexate and prednisolone and those receiving prednisolone alone (HR: 0.98; 95% CI 0.62 – 1.54; P=0.92). Twenty (14.6%) participants were lost to follow up.

**Figure 2:**
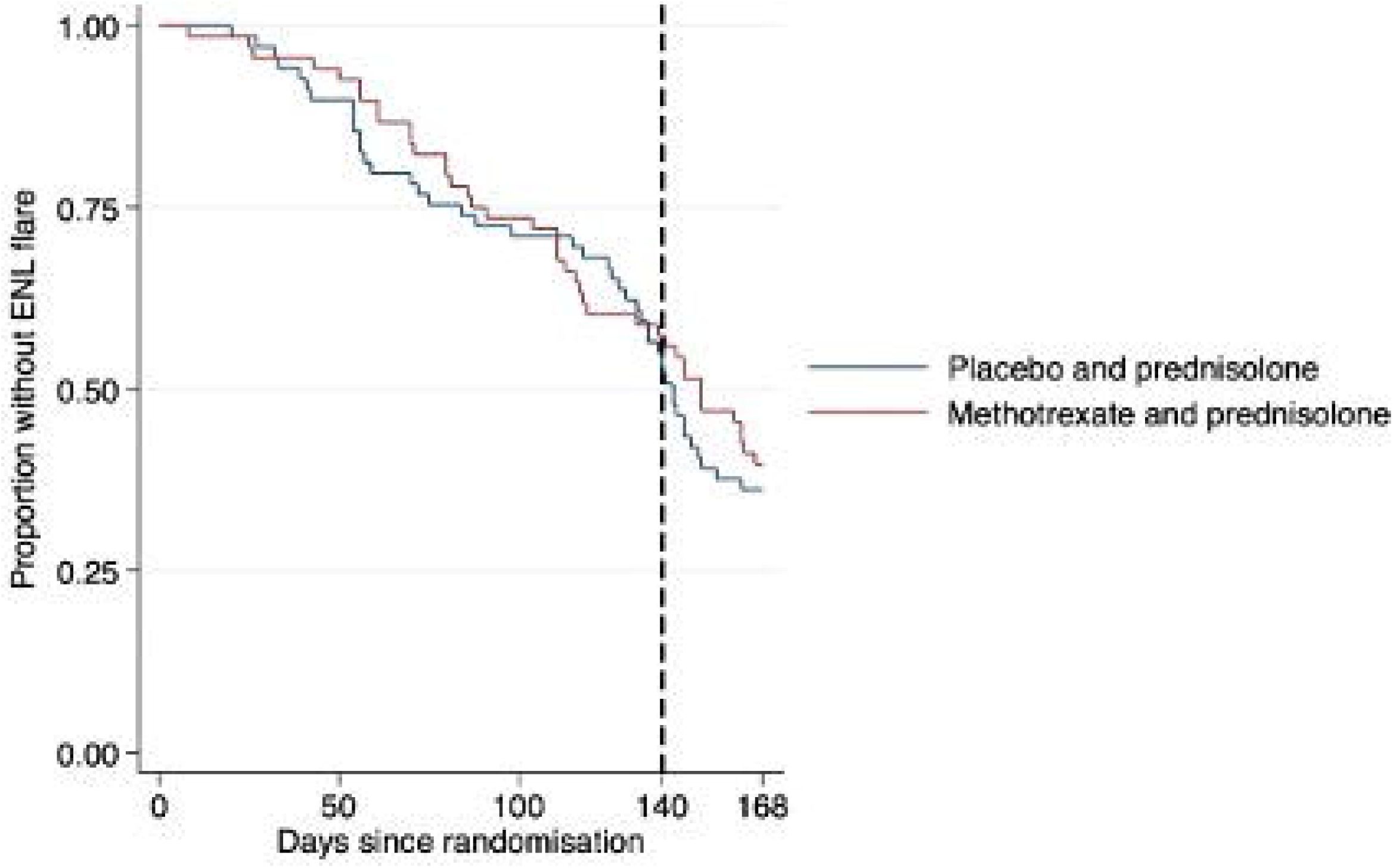
Time to first ENL flare up to 24 weeks: the Kaplan-Meier estimates of time to first ENL flare requiring additional prednisolone in the intention-to-treat population, comparing participants receiving methotrexate (MTX) and prednisolone with participants receiving placebo and prednisolone. The broken vertical line indicates the end of the prednisolone component of the intervention.

By 48 weeks, 102 of 137 participants (74.5%) had experienced at least one ENL flare requiring additional prednisolone. This occurred in 49 of 68 (72.1%) in the methotrexate and prednisolone group and 53 of 69 participants (76,8%) in the placebo and prednisolone group. The Kaplan-Meier estimate of remaining free from ENL-related additional prednisolone at 48 weeks was 27.9% (95%CI 17.9-38.9) in the methotrexate group and 23.2% (95% CI 14.1-33.6) in the prednisolone group (figure 3). In the adjusted Cox proportional hazards model, there was no evidence of a difference in the hazard of experiencing a first ENL flare requiring additional prednisolone between participants receiving methotrexate and prednisolone and those receiving placebo and prednisolone (HR: 0.95; 95%CI 0.62 – 1.43; P=0.79).

**Figure 3:**
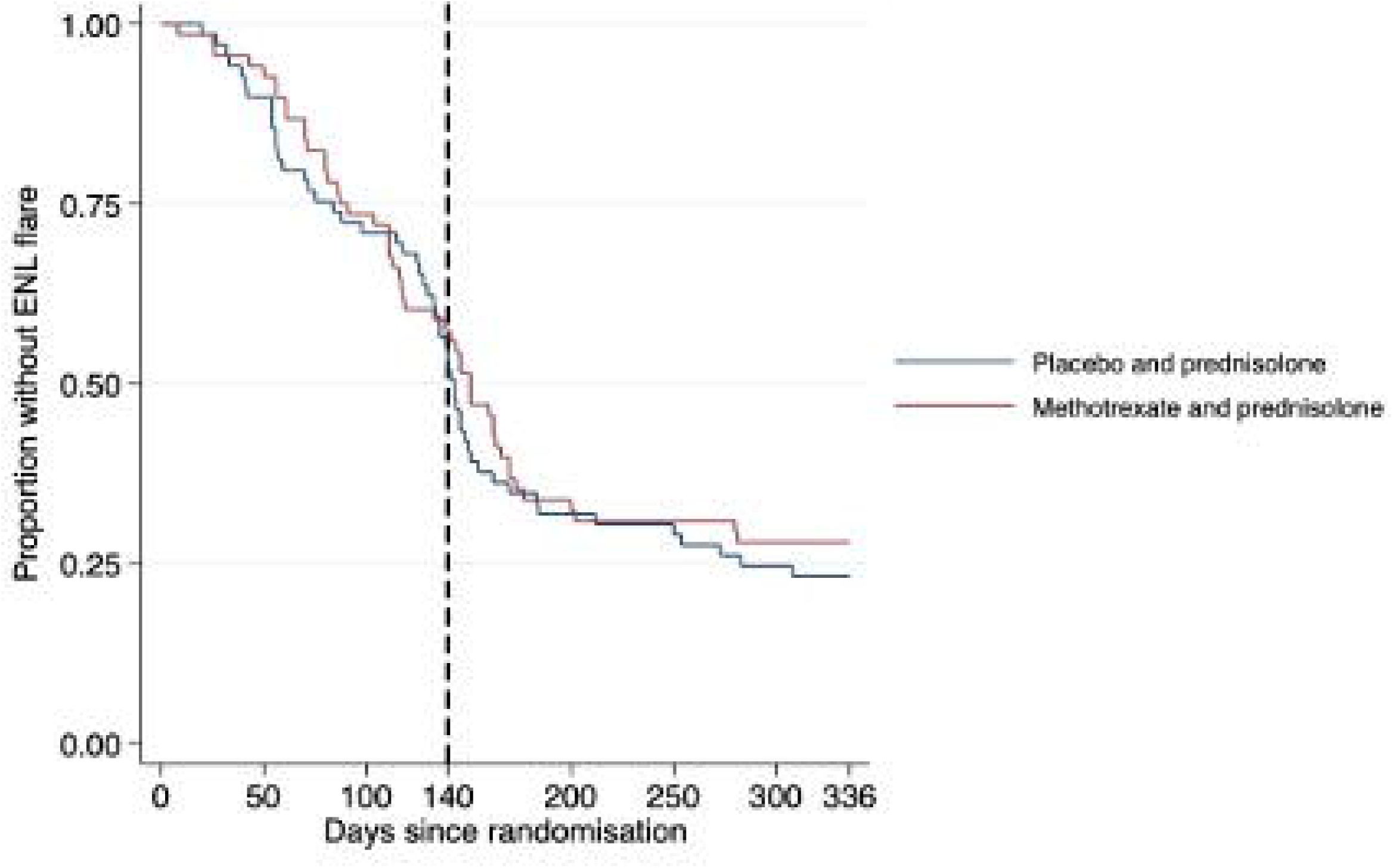
Time to first ENL flare up to 48 weeks: the Kaplan-Meier estimates of time to first ENL flare requiring additional prednisolone in the intention-to-treat population, comparing participants receiving methotrexate (MTX) and prednisolone with participants receiving placebo and prednisolone. The broken vertical line indicates the end of the prednisolone component of the intervention.

Figures 2 and 3 showed substantial overlap for time to first ENL flare between intervention groups throughout follow-up, with no clear separation before or after completion of prednisolone at Day 140.

In the per-protocol population, results were consistent with the intention to treat analysis. There was no evidence of difference between treatment arms in time to first ENL flare requiring additional prednisolone at either 24 or 48 weeks. Adjusted hazard ratios were similar, and confidence intervals overlapped those from the intention-to-treat analysis (Table 3).

**Table 3:** Hazard ratios (HR) of ENL flares forthe methotrexate and prednisolone arm compared to placebo and prednisolone arm. Intention-to-treat population and per protocol population at 24 weeks and 48 weeks

| Analysis | 24 weeks HR (95% CI) | 48 weeks HR (95% CI) |
| --- | --- | --- |
| Intention-to-treat -unadjusted | 0.89 (0.58-1.36) | 0.88 (0.6-1.3) |
| Intention-to-treat - adjusted | 0.98 (0.62–1.54) | 0.95 (0.62–1.43) |
| Per protocol -unadjusted | 0.97 (0.64-1.48) | 0.99 (0.68-1.47) |
| Per protocol - adjusted | 1.07 (0.69–1.66) | 1.06 (0.72–1.56) |

In an exploratory Cox model comparing methotrexate and prednisolone with prednisolone alone within ENL subgroups (acute ENL and recurrent/chronic ENL), methotrexate and prednisolone were associated with a higher hazard of first ENL flare requiring additional prednisolone at 24 weeks (HR:2.2; 95%CI 0.78 – 6.21). A similar pattern was observed at 48 weeks (HR:1.44; 95% 0.66 – 3.15), with no statistically significant difference between treatment groups.

### Secondary outcomes

#### Change in EESS scores from enrolment to the first flare of ENL

Amongst participants who experienced an ENL flare, EESS scores at the time of first flare were lower than at enrolment in both groups (Table 4). Overall, the median EESS score decreased from 14 (IQR 12-17) at enrolment to 11 (IQR 9-14) at first flare, corresponding to a median change of – 3 points.

**Table 4:** Change in ENLIST ENL Severity Scale score from enrolment to first ENL flare.

| Group |  | Enrolment EESS score median (IQR) | First-Flare EESS score median (IQR) | Median change |
| --- | --- | --- | --- | --- |
| **All participants at first flare (n=103)** |  | **14 (12-17)** | **11 (9-14)** | **-3** |
| **Methotrexate and prednisolone** | **All** | **14 (12-17)** | **12 (9-14)** | **-3** |
|  | **Acute ENL** | **14 (12-16)** | **12 (9-14)** | **-2** |
|  | **Recurrent or chronic ENL** | **15 (13-17)** | **12 (10-15)** | **-3** |
| **Placebo and prednisolone** | **All** | **15 (12-17)** | **12 (9-14)** | **-3** |
|  | **Acute ENL** | **14 (12-16)** | **11 (9-14)** | **-3** |

|  | Recurrent or chronic ENL | 15 (13-17) | 12 (10-14) | -3 |
| --- | --- | --- | --- | --- |

Similar reductions in EESS scores were observed across ENL subtypes in both intervention arms. Amongst Individuals with acute ENL, the median score change was -2 points in the methotrexate and prednisolone group and -3 points in the prednisolone group. Amongst Individuals with recurrent or chronic ENL, the median change was -3 points in both intervention groups.

Figure 4 shows the change in EESS score between enrolment and the first flare for all participants and stratified by intervention group and ENL type. Negative values indicate lower EESS scores at the time of first flare compared with enrolment. Boxes represent the interquartile range, the horizontal lines indicate the median, and whiskers show the range.

**Figure 4:**
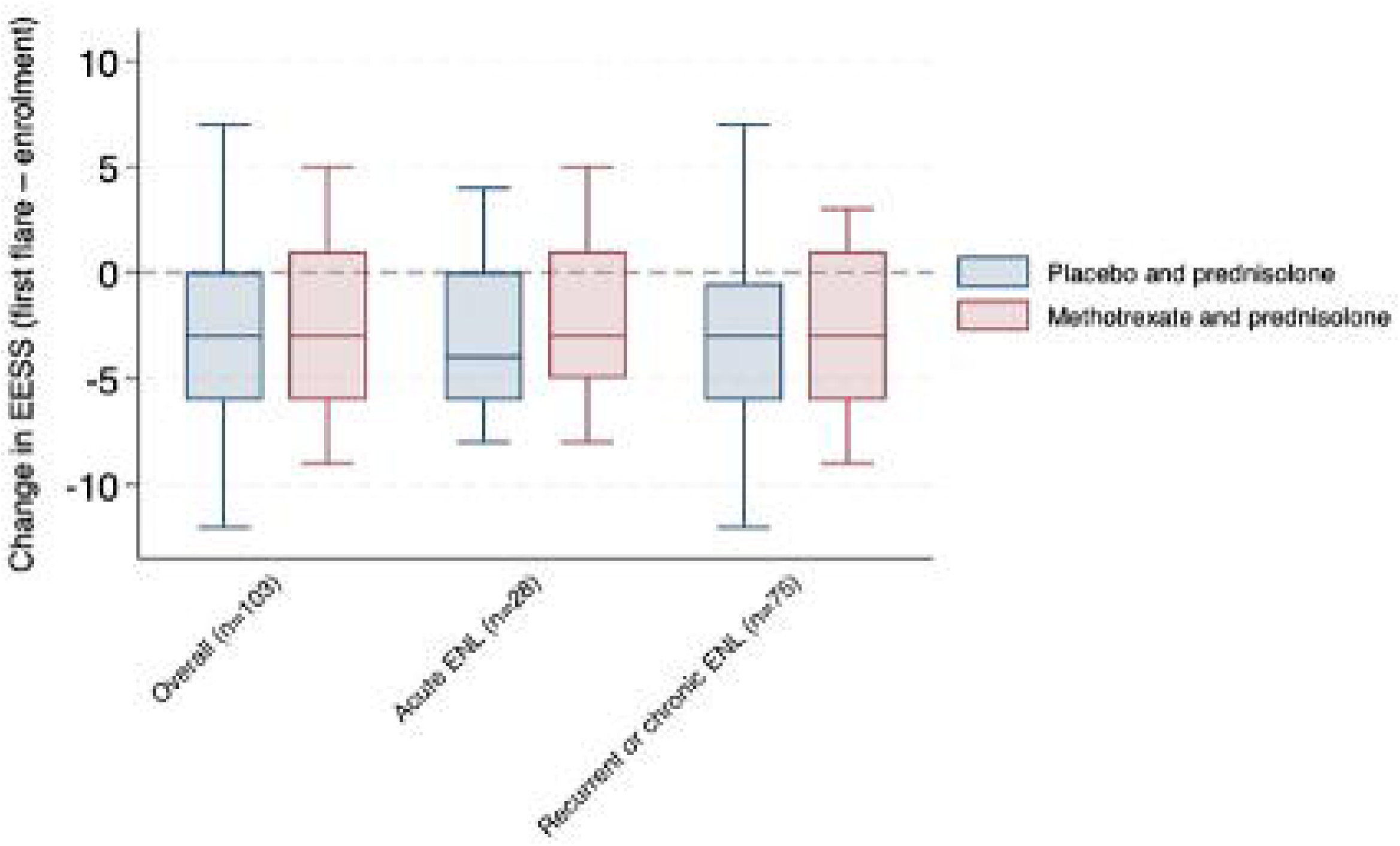
Change in ENLIST ENL Severity Scale score from enrolment to first ENL flare requiring additional prednisolone

Using median (quartile) regression, there was no evidence of a difference in the change in EESS scores from enrolment to first flare between methotrexate and prednisolone and prednisolone alone within ENL types. The estimated difference in the median change (methotrexate and prednisolone score minus prednisolone alone score) was 1 point (95%CI - 3.6 to 5.6) in acute ENL and zero (95%CI -2.7 to 2.7) in recurrent or chronic ENL.

#### Requirement for additional prednisolone at 60 weeks

One participant had their first ENL at flare after week 48, so by end of follow-up at 60 weeks, 103 of 137 participants (75.2%) had required additional prednisolone for ENL, while 34 participants (24.8%) had not. The proportion of participants requiring additional prednisolone was similar between treatment arms, occurring in 49 of 68 participants (72.1%) in the methotrexate and prednisolone arm and 54 of 69 participants (78.3%) in the prednisolone arm.

In time-to-event analyses adjusted for ENL subtype, there was no evidence of a difference between treatment arms in time to first requirement for additional prednisolone by 60 weeks. Amongst participants with acute ENL, the hazard ratio for methotrexate and prednisolone compared with prednisolone alone was 1.52 (95% CI 0.70 to 3.28; p = 0.29). Amongst those with recurrent or chronic ENL, the corresponding hazard ratio was 0.87 (95% CI 0.57 to 1.34; p = 0.54). These findings indicate no statistically significant difference between methotrexate and prednisolone and prednisolone alone in the requirement for additional prednisolone during extended follow-up.

#### Number of flares of ENL requiring additional prednisolone

Among the 103 participants who experienced at least one ENL flare requiring additional prednisolone during the study, the number of flares per participant was similar between treatment groups. Participants allocated to the methotrexate and prednisolone arm (n = 49) and the placebo and prednisolone arm (n = 54) each experienced a median of three flares requiring additional prednisolone. The interquartile range (IQR) was wider in the prednisolone alone arm (IQR 3; range 1–8) than in the methotrexate arm (IQR 1; range 1–7), but there was no evidence of a difference in flare burden between the treatment groups (Mann–Whitney U test, p = 0.56).

When stratified by ENL type at enrolment, participants with acute ENL (n = 28) and those with recurrent or chronic ENL (n = 75) also experienced a similar number of flares requiring additional prednisolone. Both groups had a median of three flares during follow-up; the IQR was 1 (range 1–5) among participants with acute ENL and 2 (range 1–8) among those with recurrent or chronic ENL. There was no statistically significant difference in flare burden between ENL subtypes (Mann–Whitney U test, p = 0.68). Table 5 summarises the number of participants who experienced one or more ENL flares. Overall, these findings indicate that neither methotrexate nor ENL subtype at enrolment was associated with a reduction in the frequency of ENL flares requiring additional prednisolone.

**Table 5:**
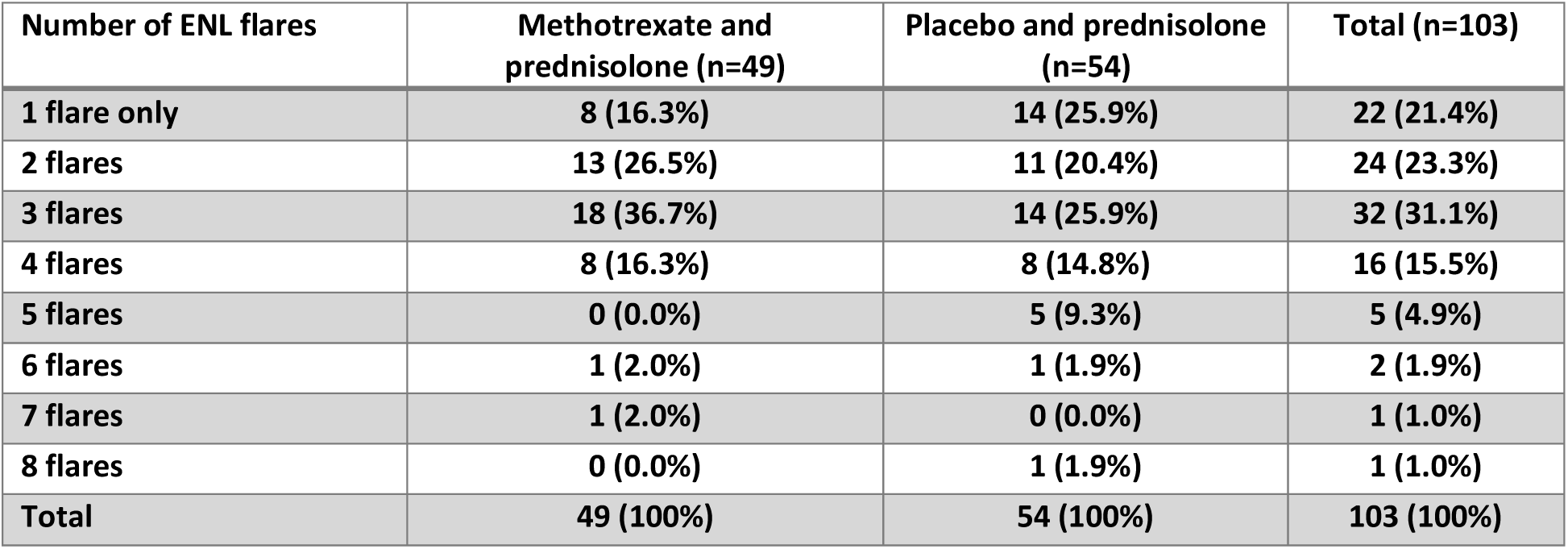
Number of participants experiencing one or more ENL flares.

| Number of ENL flares | Methotrexate and prednisolone (n=49) | Placebo and prednisolone (n=54) | Total (n=103) |
| --- | --- | --- | --- |
| 1 flare only | 8 (16.3%) | 14 (25.9%) | 22 (21.4%) |
| 2 flares | 13 (26.5%) | 11 (20.4%) | 24 (23.3%) |
| 3 flares | 18 (36.7%) | 14 (25.9%) | 32 (31.1%) |
| 4 flares | 8 (16.3%) | 8 (14.8%) | 16 (15.5%) |
| 5 flares | 0 (0.0%) | 5 (9.3%) | 5 (4.9%) |
| 6 flares | 1 (2.0%) | 1 (1.9%) | 2 (1.9%) |
| 7 flares | 1 (2.0%) | 0 (0.0%) | 1 (1.0%) |
| 8 flares | 0 (0.0%) | 1 (1.9%) | 1 (1.0%) |
| Total | 49 (100%) | 54 (100%) | 103 (100%) |

#### Severity of flares of ENL

Amongst 103 participants who experienced at least one ENL flare, a total of 282 ENL flares were recorded during follow-up. Of these, 179 flares represented second or subsequent flares and were included in analyses of change in severity between consecutive flares. The number of participants experiencing multiple flares declined progressively with increasing flare order. 81 participants had two or more flares, 57 three or more, and substantially fewer with higher-order flares.

Across all participants and flare transitions, the median change in EESS score between consecutive flares was 0 (IQR 5; range −9 to +11), indicating no consistent overall worsening or improvement in flare severity over time. When examined by flare order, the median change in EESS score was 0 for the second flare compared with the first (median 0, IQR 5), and modestly positive for the third and fourth flares compared with the preceding flare (median +1 for both transitions). Higher-order transitions were infrequent and showed variable changes in severity, with small numbers precluding reliable interpretation.

When stratified by treatment arm, patterns of change in flare severity were broadly similar between participants receiving placebo and those receiving methotrexate. For the comparison of second versus first flares, the median change in EESS score was 0 in both arms, with no evidence of a difference between groups (Mann–Whitney U test, p = 0.71). Likewise, no statistically significant differences between treatment arms were observed for the third versus second flare (p = 0.10) or the fourth versus third flare (p = 0.42). These findings indicate that methotrexate and prednisolone were not associated with a consistent reduction or increase in the severity of successive ENL flares compared with prednisolone alone.

Analyses stratified by ENL type at enrolment showed similar overall patterns, although some differences were observed in early flare transitions. For the second versus first flare, participants with acute ENL tended to experience a lower median change in EESS score compared with those with recurrent or chronic ENL (median −1 vs +1), although this difference did not reach statistical significance (p = 0.09). For the third versus second flare, participants with acute ENL experienced a greater median increase in EESS score than those with recurrent or chronic ENL (median +3 vs 0), and this difference was statistically significant (p = 0.02). No differences in change in severity between ENL subtypes were observed for later flare transitions, although sample sizes were small. Figure 5. shows the comparison between EESS scores for a flare and the flare that preceded it.

**Figure 5:**
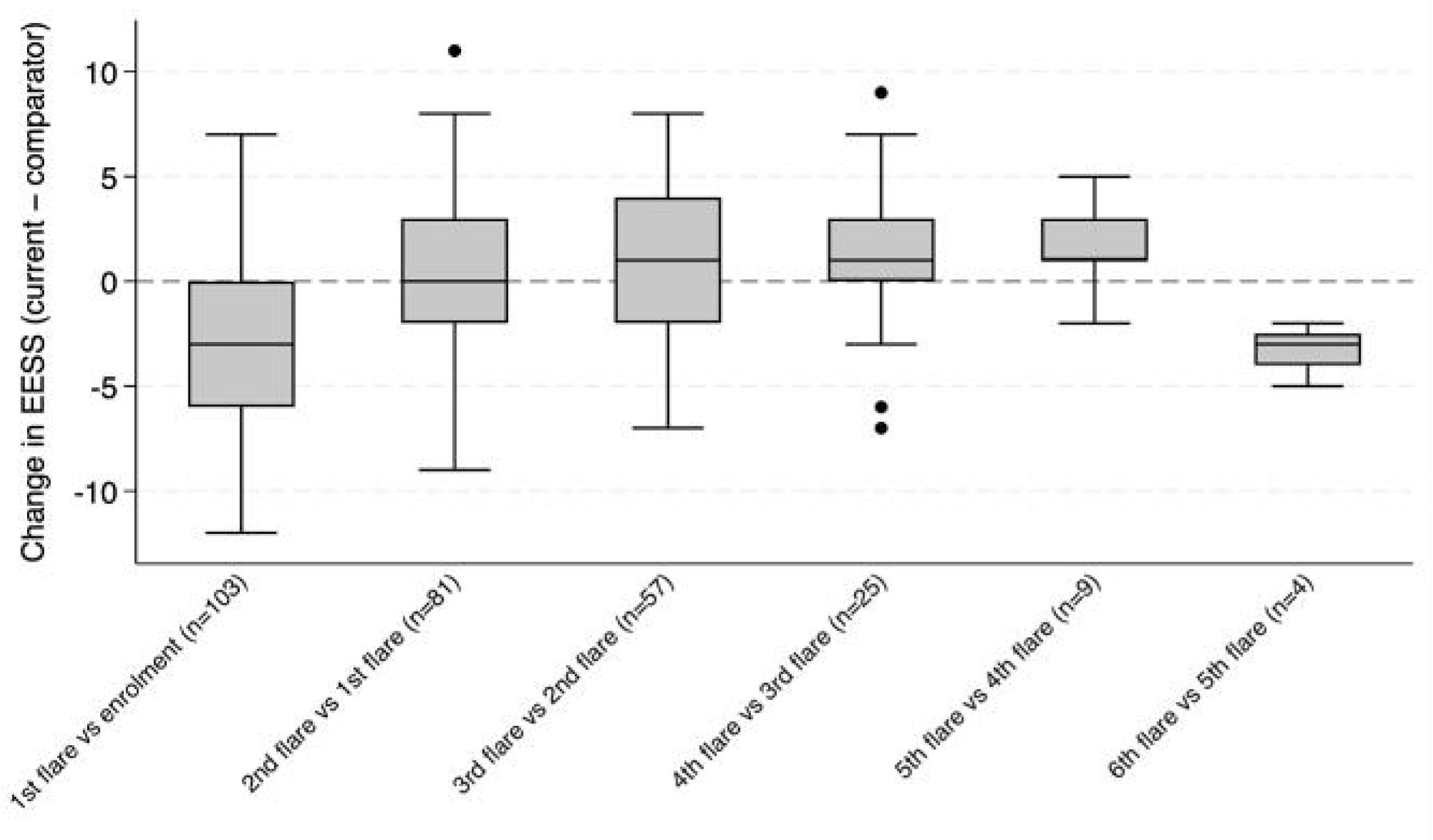
Change in ENLIST ENL Severity Scale scores between consecutive flares X-axis: consecutive ENL flare comparison. Boxes show median and interquartile range; the dashed line indicates no change in severity

Overall, these analyses suggest that the severity of ENL flares does not show a consistent pattern of progressive worsening or improvement across successive flares. While some variability in severity between consecutive flares was observed, particularly in early flare transitions, these changes were not consistently associated with treatment allocation. Differences by ENL subtype were limited to early flare transitions and should be interpreted cautiously given the declining number of participants experiencing higher-order flares.

#### Time to first flare after enrolment

The median time to first ENL flare, derived from the Kaplan-Meier estimates, was 117 days in the methotrexate and prednisolone group and 134 days in the placebo and prednisolone group. Consistent with the survival analysis presented in Figures 2 and 3, there was no statistically significant difference between intervention groups in time to first ENL flare.

#### Health-related quality of life

##### Dermatology Life Quality Index

At enrolment, HRQoL impairment was severe and similar in both groups, with a median DLQI of 19 in the methotrexate and prednisolone group (IQR-13-20) and 19 in the placebo and prednisolone group (IQR 15-21), corresponding to a very large effect on quality of life.

By week 48, DLQI scores improved substantially in both groups, with median scores of 1 (IQR 0-8) in the methotrexate and prednisolone group and 3 (IQR 0-8) in the placebo and prednisolone group. At week 60, median DLQI scores were 2 in both groups (methotrexate and prednisolone IQR 0-6.5; placebo and prednisolone IQR 0-5), indicating minimal residual impact on quality of life. Improvements in DLQI scores were observed in both treatment arms across acute and recurrent/chronic ENL (Figure 6). In participants with acute ENL, median DLQI scores decreased substantially from enrolment to weeks 48 and 60 in both the methotrexate and placebo groups. Similar patterns of improvement were seen among participants with recurrent or chronic ENL. A 4-point change in DLQI change in DLQI is considered clinically meaningful. There was no evidence of a differential treatment effect by ENL type, with overlapping distributions and no clinically meaningful differences between treatment arms at either follow-up time point.

**Figure 6:**
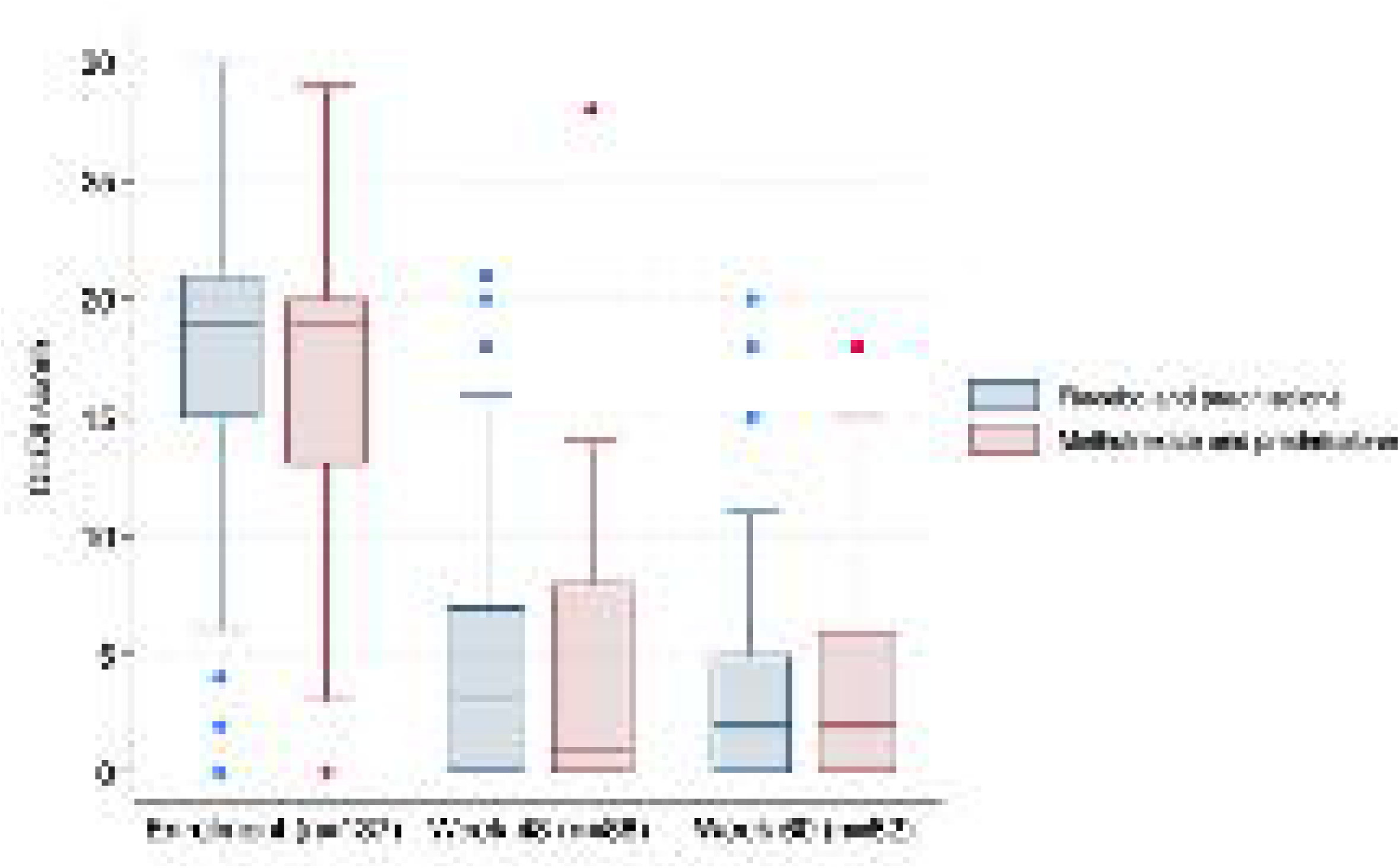
Dermatology Life Quality Index at enrolment, Week 48 and Week 60. Boxes show median and interquartile range

In adjusted analyses, there was no evidence of a difference between treatment groups at either time point. At week 48, the adjusted mean difference (methotrexate and prednisolone vs placebo and prednisolone) was -1 point (95%CI -3.5 to 1.5; p=0.43), and at week 60 it was -0.2 points (95%CI -2.4 to 2.1; p=0.89) (Table 6). These differences were below the stablished 4-point threshold for clinically meaningful change.

**Table 6:** Change in Dermatology Life Quality Index score from enrolment to weeks 48 and 60.

| Assessment | Methotrexate and prednisolone median (IQR) | Placebo and prednisolone median (IQR) | Adjusted mean difference† (95% CI) | p-value |
| --- | --- | --- | --- | --- |
| Enrolment | 19 (13–20) | 19 (15–21) | — | — |
| Week 48 | 1 (0–8) | 3 (0–7) | -1.0 (-3.5 to 1.5) | 0.43 |
| Week 60 | 2 (0–6.5) | 2 (0–5) | -0.2 (-2.4 to 2.1) | 0.89 |

†Adjusted mean differences estimated using analysis of covariance (ANCOVA), adjusting for enrolment Dermatology Life Quality Index score. Negative values favour methotrexate.

##### 36-Item Short Form Survey Instrument – SF-36

Across all SF-36 domains, participants in both treatment groups experienced marked improvements from enrolment to weeks 48 and 60. Median scores increased substantially in all domains. Analyses of SF-36 outcomes excluded the five participants recruited in Ethiopia, as the SF-36 questionnaire version used in the trial has not been formally validated in Amharic. Paired analyses demonstrated statically significant improvements from enrolment to week 48 and week 60 across all SF-36 domains (all p<0.001). Additional information of SF-36 results is available in the Supplementary File 2.

Adjusted analyses showed no consistent evidence of a treatment effect favouring methotrexate and prednisolone at either week 48 or week 60. Although point estimates favoured methotrexate in some domains (physical role and emotional role) at week 48, confidence intervals were wide and included the null, and these effects were not sustained at week 60. Most adjusted mean differences were below the commonly accepted 5–10-point threshold for a clinically meaningful change. Additional information of SF-36 analyses is available in Supplementary File 2.

#### Adverse events and serious adverse events

Across follow-up, 111 non-serious adverse event (AE) reports were recorded, of which 105 (94.6%) were judged to be related to trial treatment (methotrexate and/or prednisolone). The most frequent treatment-related AE category was liver function abnormality defined by elevation of serum transaminases (ALT and or AST) to 2 times the upper limit of normal (47/105, 44.8%), followed by anaemia (<Hb 9g/dl) (28/105, 26.7%) and thrombocytopenia (<100,000/mm^3^) (19/105, 18.1%). Less frequent treatment-related events included infection (5/105, 4.8%), metabolic events (3/105, 2.9%), musculoskeletal (low back pain and spontaneous fracture of lumbar vertebrae) events (2/105, 1.9%) and nausea and vomiting (1/105, 1.0%).

All treatment-related laboratory abnormalities were asymptomatic, and biochemical parameters improved following temporary interruption or discontinuation of the study medication.

At the participant level, 55 participants experienced at least one treatment-related non-serious adverse event during follow-up. Liver function abnormalities were the most common treatment-related adverse event, affecting 30/55 participants (54.5%), followed by anaemia (20/55, 36.4%) and thrombocytopenia (11/55, 20.0%).

When summarised descriptively by adverse event category amongst affected participants, liver function abnormalities (increased transaminases) were more frequently observed in the methotrexate arm (21/30 participants), whereas thrombocytopenia occurred predominantly in the placebo arm (10/11 participants). Anaemia was observed in both treatment groups. Participants who were receiving MDT during the trial had dapsone stopped at enrolment to minimise the risk of dapsone associated anaemia. Musculoskeletal and gastrointestinal adverse events were rare, with only one participant experiencing musculoskeletal events and one participant experiencing a gastrointestinal event across the trial.

These summaries reflect the distribution of adverse events amongst participants experiencing treatment-related non-serious adverse events and should not be interpreted as incidence estimates for the full trial population.

There were eight serious adverse events (SAEs). Five were treatment related, which were infections requiring hospitalisation and withdrawal from the trial. The SAEs were cellulitis, neck abscess, tuberculous meningitis and pulmonary tuberculosis managed according to clinical protocols and all five individuals were discharged home. The three non-treatment related SAEs were a positive pregnancy test on visit 18 and subsequently lost to follow-up and two road traffic accidents (one of which was fatal). Details of SAEs are in Supplementary File 3.

## DISCUSSION

In this international, multicentre, double-blind, randomised, placebo-controlled trial weekly oral methotrexate added to standard prednisolone treatment did not reduce the requirement for additional prednisolone in individuals with severe ENL. Methotrexate was not associated with a reduction in the occurrence of ENL flares requiring additional prednisolone, nor did it delay time to first flare. These findings were consistent across intention-to-treat and per-protocol analyses and were robust to prespecified covariate adjustment.

Of 231 individuals screened, 137 participants with severe ENL were randomised with high median EESS score at enrolment. By 24 weeks, nearly two-thirds of participants had experienced at least one flare requiring additional prednisolone increasing to three-quarters by 48 weeks, underscoring the relapsing and treatment-refractory nature of severe ENL.

Secondary outcomes were concordant with the primary analysis. Methotrexate did not reduce ENL severity at relapse, the number of flares requiring additional prednisolone, or the severity of consecutive flares. In contrast, HRQoL improved substantially from enrolment to 48 weeks and was sustained to 60 weeks in both intervention groups, as measured by DLQI and SF-36. This likely reflects the overall clinical improvement with prednisolone and close follow-up rather than a treatment-specific effect of methotrexate.

We cannot recommend the use of methotrexate for the management of severe ENL. The absence of a demonstrable benefit of methotrexate in this trial is unlikely to be explained solely by limited power. The observed differences between treatment arms were small, and even with a larger sample size, a clinically meaningful effect appears to be an unlikely outcome. Only seven double-blind randomised trials have evaluated pharmacological interventions for ENL, collectively recruiting 225 participants [34–36]. The randomised trial of methotrexate in ENL had 19 participants, no allocation concealment, only ten participants received methotrexate 7.5 mg weekly, a lower dose than used in our study. No validated outcome measure was used and there was no significant difference reported in the proportion of individuals requiring additional prednisolone.

It is possible that methotrexate 15 mg or 20 mg weekly may be relatively insufficient for treatment of severe ENL. However, this dosing regimen is consistent with established dermatological guidance for inflammatory skin disease and falls within the recommended therapeutic range [27]. It is also comparable to a large cohort of methotrexate use for leprosy reactions from France [37], in which a median dose of 20mg/week was administered.

ENL is a complex, multisystem inflammatory condition [12] and the immunomodulatory mechanism of low-dose methotrexate may not be sufficiently potent or lack the necessary specificity to modify the pathogenic inflammatory pathways in severe ENL including TNF-α associated inflammation [24,38]. Methotrexate is effective in other inflammatory conditions associated with increased levels of TNF-α such as psoriasis and psoriatic arthritis [27, 39]. Anti-TNF-α blockers used in psoriasis have been reported to be effective in ENL [40–44]. Thalidomide which is effective in ENL is not effective in other TNF-α mediated conditions such as toxic epidermal necrolysis [45,46]. These paradoxes highlight the need for improved understanding of the complex pathogenetic mechanisms involved in ENL. Despite the effectiveness of thalidomide, it is restricted or prohibited in many endemic countries due to teratogenicity, and even if available is often not affordable for people with ENL [47].

Methotrexate may still have a role in Type 1 leprosy reactions (T1R) and neuritis, for which prolonged corticosteroid therapy remains the main treatment. Evidence supporting methotrexate in T1R is currently derived largely from retrospective and uncontrolled studies [38,48]. Interestingly T1R are associated with increased tissue levels of TNF-α but do not respond to thalidomide [49,50].

Methotrexate was generally well tolerated. There was no excess of serious or clinically significant adverse events. These findings are consistent with the established safety profile of low-dose methotrexate [51–54] and demonstrate that it can be safely administered with appropriate monitoring and folic acid supplementation in leprosy-endemic settings.

We have conducted the largest double-blind, randomised, placebo-controlled trial in ENL to date. The multicentre design, with clinically meaningful outcome measure and clear criteria for additional prednisolone and use of validated outcomes measures. The limitations include failure to reach the original target sample size, requiring combined analysis of participants with acute and recurrent or chronic ENL. The final analyses were interpreted with appropriate caution, focusing on the magnitude and direction of effects rather than statistical significance alone.

Future treatment trials for ENL should incorporate many of the elements developed for MaPs in ENL, such as standardised inclusion criteria and a clinically relevant outcome measure. There is a need for evidence-based and consensus on ENL classifications to inform inclusion criteria and analysis.

## CONCLUSION

ENL remains a severe, neglected complication of leprosy with very limited alternatives to prolonged, high dose oral corticosteroid therapy. The morbidity and mortality associated with severe ENL requires continued efforts to improve the management by identifying safe, alternative immunomodulatory or corticosteroid-sparing drugs that are scalable, and deliverable in leprosy-endemic settings.

## SUPPORT

This work was supported by The Hospital and Homes of St. Giles, grant number ITCRZM25 and Leprosy Research Initiative, Turing Foundation and plan:g under LRI grant number 704.16.71

## Supporting information

Statistical Analysis Plan

Adverse Events tables

## Data Availability

De-identified participant data that support the findings of this study will be made available through the London School of Hygiene & Tropical Medicine Data Compass repository following a two-year embargo period after publication, as additional analyses are ongoing. Data will then be available upon reasonable request and subject to institutional and ethical approvals.

## ACKNOWLEDGEMENTS

We would like to express our gratitude to the people with lived experience of leprosy who participated in, or considered participating in, this study. Their time and commitment made this research possible.

We thank the members of the independent Data and Safety Monitoring Board: Emily Nightingale (statistician), Dr Pepy Dwi Endraswari, Dr Elinor Moore (Chair).

We are grateful to Professor Harparkash Kaur for conducting independent drug quality control testing, and to the Cipla Foundation for their support.

We thank the clinical and research staff at all participating centres for their dedication to recruitment, follow-up, and patient care.

We acknowledge the contribution of the ENLIST Group, including Marivic Balagon, C. Ruth Butlin, Milton O. Moraes (in memoriam), Jose da Costa Nery, and Anna Sales, for their continued commitment to advancing research in ENL and improving outcomes for affected communities.

## DATA AVAILABILITY STATEMENT

The de-identified individual participant dataset underlying the results reported in this article, together with the data dictionary and statistical analysis code, will be deposited in the LSHTM Data Compass repository. The dataset will be made publicly available following a one-year embargo after publication of the article to allow completion of planned secondary analyses by the study investigators.

## AUTHOR CONTRIBUTIONS

Conceptualisation: Barbara de Barros, Diana N. J. Lockwood, Stephen L. Walker and the Erythema Nodosum Leprosum International Study Group (ENLIST)

Methodology: Barbara de Barros, Bernd Genser, Peter Nicholls, Diana N. J. Lockwood and Stephen L. Walker

Statistical Analysis: Barbara de Barros, Stephen L. Walker and Bernd Genser

Investigation: Farha Sultana, Anju Wakade, Bhagyashree Bhame, Bishwanath Acharya, Abdulnaser Hamza, Alemtsehay Getachew, Medhi Denisa Alinda, M. Yulianto Listiawan, Shimelis N Doni, Deana A. Hagge, Indra Napit, Mahesh Shah, Vivek V. Pai, Neeta Maximus, Joydeepa Darlong

Data Curation: Barbara de Barros, Bernd Genser, Peter Nicholls

Project Administration: Barbara de Barros

Supervision: Diana N. J. Lockwood and Stephen L. Walker

Writing – Original Draft Preparation: Barbara de Barros

Writing – Review & Editing: All authors

Funding Acquisition: Diana N. J. Lockwood, Stephen L. Walker

