## Supplementary material for "Methotrexate and Prednisolone compared to placebo and prednisolone in the treatment of Erythema Nodosum Leprosum - an international multicentre, double-blind randomised controlled clinical trial - MaPs in ENL": Statistical Analysis Plan

| | |
|---|---|
| **Name: Stephen Walker** | **Role: Chief Investigator** |
| **Name: Barbara de Barros** | **Role: Study Manager** |
| **Name: Peter Nicholls and Barbara de Barros** | **Role: Data curator** |
| **Name: Peter Nicholls and Bernd Genser** | **Role: Statistician** |
| **Version: 2** | **Date: 14/11/2025** |

NCT 03775460

### 1. Introduction

This revised Statistical Analysis Plan (SAP) updates the original, pre-registered plan for the MaPs trial. Due to challenges in recruitment and feasibility constraints, the final sample size was lower than originally anticipated. Before unblinding, the ENLIST group (trial steering committee), the study statistician, chief investigator and study manager approved combining acute and chronic/recurrent ENL into a single analysis population. This SAP details the rationale for these changes and the updated statistical methods to be used.

### 2. Rationale for SAP Revision

The original SAP specified separate analyses for acute and chronic/recurrent ENL populations. However, despite extended recruitment and multiple study sites, total enrolment remained below the planned target. Under-recruitment is common in clinical trials, especially in rare conditions, and does not invalidate randomisation, provided analyses are pre-specified and appropriately adapted.

To maximise scientific value and preserve randomisation integrity, acute and chronic/recurrent ENL participants will now be analysed as a combined primary population. ENL type will be included as an adjustment and subgroup variable. This modification does not change the original research question[1–3].

#### Justification for the Ad Hoc Combined Analysis

- **Rarity of ENL and recruitment feasibility:** ENL is a rare complication of leprosy, and eligible cases remained limited despite opening multiple international centres and extending recruitment. Under-recruitment is widely recognised in trials of rare diseases.
- **Preservation of randomisation:** Combining ENL types maintains the validity of randomisation while maximising statistical information. No conceptual conflict is introduced, as acute and chronic/recurrent ENL share pathophysiological mechanisms and treatment pathways.

- **Estimation-focused inference:** Given the reduced sample size, emphasis will be placed on treatment-effect estimation with confidence intervals rather than hypothesis testing, consistent with modern methodological recommendations.
- **Largest ENL trial to date:** Even with fewer participants than planned, MaPs remains the largest randomised evaluation of ENL treatment ever conducted; combining groups ensures that the data yield clinically meaningful estimates.

This modification does not change the underlying research question. ENL type will be included as an adjustment variable and examined in pre-specified subgroup analyses.

**3. Analysis Populations**

- **Intention-to-treat (ITT):** All randomised participants, analysed by allocation.
- **Per-protocol (PP):** ITT participants without major protocol deviations.
- **Safety population:** All participants receiving at least one dose of study medication.

**4. Primary Outcomes**

**Primary outcomes:**

1. Requirement for additional prednisolone between randomisation and week 24.

2. Requirement for additional prednisolone between randomisation and week 48.

The difference in probability of receiving any additional prednisolone between treatment arms in the overall ENL population.

**5. Statistical Methods for Primary Outcomes**

- **Unadjusted analyses**
  - Kaplan–Meier curves by treatment arm.
  - Kaplan-Meier curves stratified by arm × ENL type (acute vs recurrent/chronic).
  - Log-rank test used descriptively.

- **Adjusted analyses:**
  - Cox model includes the following covariates regardless of baseline balance: treatment, ENL type, sex, age (categorical), BMI, severity score.

- **Missing data:** MMRM as primary approach; LOCF and best/worst-case as sensitivity analyses.
- **Per-protocol:** Repetition of primary models.

**6. Secondary Outcomes**

- **Continuous outcomes** (e.g., EESS, DLQI, SF-36): Mixed-effects linear models.
- **Count outcomes** (e.g., number of ENL flares): Poisson/negative binomial models.

- **Time-to-event outcomes:** Kaplan–Meier curves and Cox models. These analyses will be interpreted cautiously given reduced sample size.

**7. Subgroup and Exploratory Analyses**

- Subgroups: acute vs chronic/recurrent ENL, centre, sex, baseline severity.
- Interaction tests will be exploratory due to limited power.
- Forest plots will present subgroup estimates.

**8. Interpretation Considering Reduced Sample Size**

Lower-than-planned recruitment reduces statistical power, especially for subgroup analyses. The trial will focus on effect sizes and CIs rather than hypothesis testing. Despite limitations, MaPs remains the largest randomised ENL trial to date, and the combined analysis yields the most informative estimates currently available.

**9. Deviations from Original SAP**

- Combining acute and chronic/recurrent ENL populations.
- Revising analysis population definitions.
- Adopting estimation-focused interpretation. All deviations were decided prior to unblinding and approved by the ENLIST group and study statistician.

**10. Software**

Analyses will be conducted using Stata/SE 17.
