## Supplemental Data 1 for "Methotrexate and Prednisolone compared to placebo and prednisolone in the treatment of Erythema Nodosum Leprosum - an international multicentre, double-blind randomised controlled clinical trial - MaPs in ENL"

**Supplementary file 2 – Adjusted Cox proportional hazard models – Graphs and tables HRQoL**
Title: Methotrexate and Prednisolone compared to prednisolone alone in Erythema Nodosum Leprosum - an international multicentre, double-blind randomised controlled clinical trial MaPs in ENL

This supplementary file presents additional analyses of health-related quality of life (HRQoL) assessed using the 36-Item Short Form Health Survey (SF-36) and the Dermatology Life Quality Index (DLQI) in the Methotrexate and Prednisolone study in erythema nodosum leprosum (MaPs). Summary scores over time and adjusted between-group comparisons are provided to support the main trial findings.

DLQI and SF-36 assessments were performed at enrolment and at weeks 48 and 60. Higher SF-36 scores indicate better health status, while lower DLQI scores indicate less impairment in dermatology-related quality of life.

Analyses of SF-36 excluded five participants recruited in Ethiopia, as the SF-36 questionnaire version used in this study has not been validated in Amharic.

**Figure 1** presents box plots of DLQI scores by type of ENL (acute and recurrent or chronic ENL) over the course of follow-up. At enrolment, DLQI scores indicated marked impairment in dermatology-related quality of life in both groups. DLQI scores improved over time in both groups, with distributions narrowing during follow-up, indicating reduced variability and overall improvement in skin-related quality of life.

No clear or sustained separation between groups was observed in DLQI distributions over follow-up.

**SF-36 scores over follow-up**

**Table 1** shows median (interquartile range) SF-36 domain scores at enrolment, week 48 and week 60 by treatment arm. At enrolment, participants in both groups reported substantial impairment across physical, emotional and social domains. By weeks 48 and 60, improvements were observed in both treatment arms across all SF-36 domains, including physical functioning, bodily pain, vitality and mental health. Improvements were sustained at week 60.

**Adjusted between-group comparisons (SF-36)**
**Table 2** presents adjusted mean differences in SF-36 domain scores between the methotrexate plus prednisolone group and the prednisolone-alone group at weeks 48 and 60.

Differences were estimated using analysis of covariance (ANCOVA), adjusting for enrolment SF-36 scores. Negative values indicate higher scores (better HRQoL) in the methotrexate group.

At both time points, adjusted differences between groups were small across most domains. Whilst modest differences were observed for role-physical and role-emotional domains at week 48, adjusted differences at week 60 were minimal and not statistically significant across all SF-36 domains.

A difference of 5–10 points is commonly considered clinically meaningful for SF-36 domains; most adjusted differences observed in this study were below this threshold.

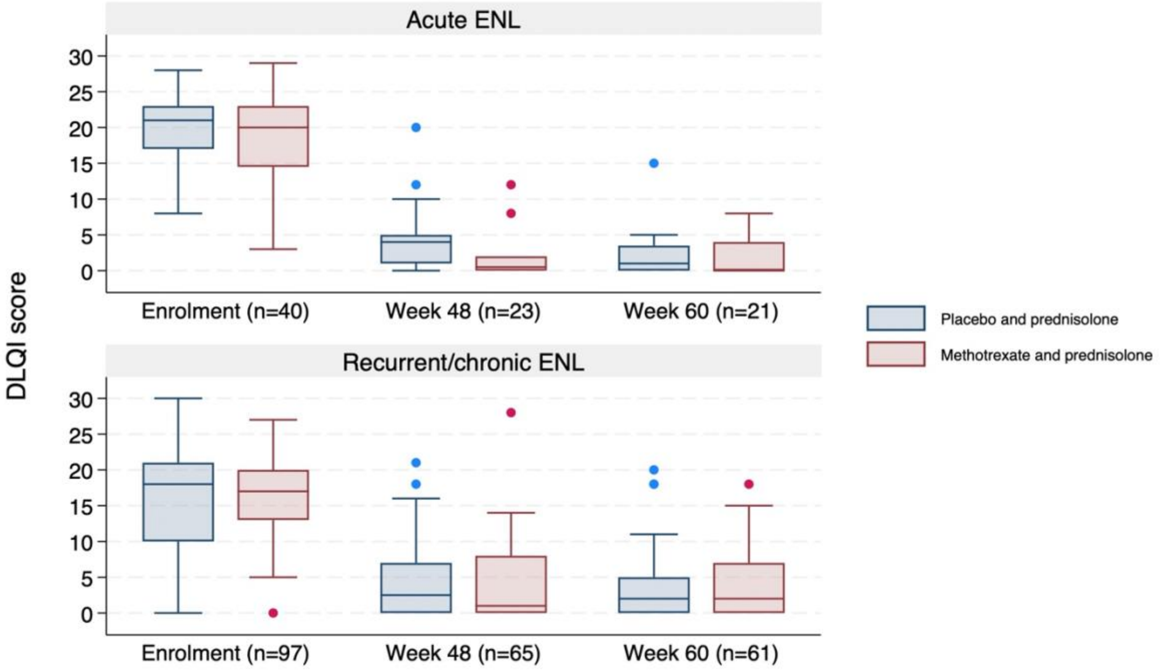


*Figure 1: DLQI over time by ENL type. Boxes show median and interquartile range*

*Table 1:SF-36 scores over trial follow up*

| | Methotrexate and prednisolone | | | Placebo and prednisolone | | |
|---|---|---|---|---|---|---|
| **SF-36 domain** | **Enrolment** | **Week 48** | **Week 60** | **Enrolment** | **Week 48** | **Week 60** |
| **Physical Functioning** | **30 (10–53.8)** | **97.5 (66.3–100)** | **95 (60–100)** | **25 (10–45)** | **100 (65–100)** | **100 (70–100)** |
| **Role Physical** | **0 (0–0)** | **100 (100–100)** | **100 (100–100)** | **0 (0–0)** | **100 (50–100)** | **100 (100–100)** |
| **Vitality** | **20 (20–35)** | **75 (66.3–80)** | **70 (65–80)** | **20 (20–35)** | **70 (55–80)** | **70 (65–75)** |
| **Mental Health** | **28 (20–43)** | **78 (69–92)** | **76 (67–92)** | **28 (20–44)** | **76 (66–86)** | **76 (72–92)** |
| **Social Functioning** | **62.5 (50–75)** | **87.5 (75–100)** | **87.5 (75–100)** | **62.5 (37.5–75)** | **75 (75–100)** | **75 (75–100)** |
| **Bodily Pain** | **32.5 (22.5–45)** | **95 (77.5–100)** | **90 (77.5–100)** | **22.5 (22.5–45)** | **87.5 (67.5–100)** | **90 (77.5–100)** |
| **General Health** | **30 (25–35)** | **45 (35–75)** | **55 (40–76.3)** | **30 (25–30)** | **55 (35–70)** | **55 (35–80)** |
| **Role Emotional** | **0 (0–0)** | **100 (100–100)** | **100 (100–100)** | **0 (0–0)** | **100 (33.3–100)** | **100 (100–100)** |

*Table 2: Adjusted differences in SF-36 domains between methotrexate and placebo at week 48 and 60*

| SF-36 domain | Week 48 adjusted mean difference† (95% CI) | p-value | Week 60 adjusted mean difference† (95% CI) | p-value |
|---|---|---|---|---|
| **Physical Functioning** | **+0.5 (−9.8 to 10.9)** | **0.92** | **−0.4 (−11.6 to 10.8)** | **0.95** |
| **Role Physical** | **+18.4 (3.4 to 33.4)** | **0.017** | **+5.9 (−9.5 to 21.3)** | **0.45** |
| **Vitality** | **+4.3 (−3.0 to 11.6)** | **0.24** | **−0.1 (−8.6 to 8.3)** | **0.97** |
| **Mental Health** | **+4.0 (−3.8 to 11.9)** | **0.31** | **−0.7 (−9.2 to 7.8)** | **0.87** |
| **Social Functioning** | **+4.2 (−3.6 to 12.0)** | **0.29** | **+0.8 (−7.5 to 9.2)** | **0.84** |
| **Bodily Pain** | **+8.2 (−1.5 to 17.9)** | **0.096** | **+0.4 (−8.5 to 9.3)** | **0.92** |
| **General Health** | **+0.4 (−10.6 to 11.4)** | **0.94** | **−0.5 (−11.4 to 10.5)** | **0.93** |
| **Role Emotional** | **+14.7 (−0.6 to 30.0)** | **0.059** | **+2.4 (−13.5 to 18.2)** | **0.77** |

*†Adjusted mean differences estimated using analysis of covariance (ANCOVA), adjusting for enrolment SF-36 score.*
*Negative values favour methotrexate.*
*A difference of 5-10 points in commonly considered clinically meaningful for SF-36 domains.*
*SF-36 analysis excluded participants recruited in Ethiopia, as the SF-36 questionnaire has not been validated in Amharic.*
