## Supplementary material for "Methotrexate and Prednisolone compared to placebo and prednisolone in the treatment of Erythema Nodosum Leprosum - an international multicentre, double-blind randomised controlled clinical trial - MaPs in ENL": Adverse Events tables

**Supplementary file 3 – Adverse Events tables**

Title: Methotrexate and Prednisolone compared to prednisolone alone in Erythema Nodosum Leprosum - an international multicentre, double-blind randomised controlled clinical trial MaPs in ENL

*Table 1: Serious treatment related adverse events*

| Serious adverse event | Outcome |
|---|---|
| **Cellulitis with infected ulcer** | **Hospitalised; treated with intravenous antibiotics and surgical debridement; improved and discharged** |
| **Abscess** | **Hospitalised; treated with intravenous antibiotics and surgical drainage; improved and discharged** |
| **Tuberculous meningitis** | **Withdrawn from study; discharged on anti-tuberculosis treatment; recovering at home** |
| **Cellulitis** | **Treated with intravenous antibiotics; improved and discharged; withdrawn from the study** |
| **Pulmonary tuberculosis** | **Treated; improved and discharged; withdrawn from the study** |
